# Long-term corticosteroid therapy for patients with severe coronavirus disease 2019 (COVID-19)

**DOI:** 10.1101/2021.08.30.21262824

**Authors:** George Calcaianu, Samuel Degoul, Thibault Payen, Bénédicte Michau, Mihaela Calcaianu, Bree Lawson, Didier Bresson, Didier Debieuvre

## Abstract

earlier and longer corticosteroid therapy with methylprednisolone could reduce the mortality and/or rate of ICU admission by 26% in patients with severe COVID-19, hospitalized in conventional medical ward.

**Background:** Low dose of dexamethasone reduced mortality in hospitalized COVID-19 patients who required respiratory support. Less is known about the efficacy of other corticosteroids in severe COVID-19 patients. This study was designed to determine if longer and earlier corticoid therapy in severe COVID-19 patients is associated with reduced mortality and/or reduced rate of ICU admission for worsening of respiratory state.

**Methods:** We performed a retrospective study with patients aged ≥ 18 years, with epidemiological and/or radiological suspected COVID-19, hospitalized in a regional hospital GHRMSA Mulhouse, France. Twenty-three patients received methylprednisolone (MP) as compassionate use, 1 mg/kg/day for seven days followed by prednisolone at a gradually reduced dosage, for 4 to 6 weeks. MP was started one week after COVID-19 symptoms first appeared. The primary composite outcome was mortality and/or ICU admission during hospitalisation.

**Results:** Between March 14^th^ to June 5^th^ 2020, 255 patients were screened, 181 were included, and 92 were analysed, 23 patients treated with MP and 69 received standard care. SARS-CoV2 infection was confirmed by reverse transcriptase polymerase chain reaction (RT-PCR) in 82.3%. The overall composite outcome was higher in the control group: 42/69 patients (60.9%) versus 8/23 (34.8%) in the interventional group (p= 0.018). The correction of lymphopenia between days 1 to 7 was associated with better outcome (p=0.006).

**Conclusion:** These results suggest that earlier and longer corticosteroid therapy with methylprednisolone could reduce the mortality and/or rate of ICU admission in patients with severe COVID-19, hospitalized in conventional medical ward.

---

Coronavirus Disease 2019 (COVID-19) was first reported in late December 2019, in Wuhan, China. Currently, there are over 213□million confirmed cases worldwide and more than 4.4 million deaths due to SARS-CoV2 infection. The Grand-Est region was one the most affected by this pandemic in France, with more than 3700 deaths. The most common cause of death in COVID-19 patients is respiratory failure with acute respiratory distress syndrome (ARDS). Several studies reported high mortality rate among in-hospital COVID-19 patients (1,2).

The pathophysiology of severe COVID-19 is characterized by acute extensive pneumonia complicated by inflammatory infiltrates, micro thrombosis and finally diffuse alveolar damage. The lung injury is not only associated to virus-induced injury, but also to the immune response triggered by COVID-19 with activation of pro-inflammatory cytokines (3–5). It has been observed that a subgroup of COVID-19 patients have markedly elevated inflammatory markers such as C-reactive protein, ferritin and interleukin 6 (6–8). Systemic corticosteroids have an important anti-inflammatory effect and are frequently used as additional treatment for viral pneumonia.

Results from a large randomized controlled trial (RCT) (RECOVERY Trial), using dexamethasone 6mg once daily for 10 days, found that the absolute risk of death was reduced by 12,1%, mostly in critically ill COVID-19 patients (9).

However, the question about the efficacy of other systemic corticosteroids such as methylprednisolone (MP), a common steroid used in clinical practice, is still debated (10–12). However, some data are consistent with the efficacy of corticosteroids independent of the formulation (13). The optimal dose and duration of treatment are still questionable.

The recently published data from the METCOVID randomized controlled trial showed that the use of MP at 1mg/kg/day, during 5 days, in critical COVID-19 patients did not improve prognosis (10). The authors of this study hypothesized that a possible explanation of this result is the shorter duration of corticoid treatment and a late start of MP treatment in the evolution of disease. A recent review of corticosteroid therapy studies showed a delayed viral clearance for SARS-CoV1 and MERS-CoV infections. It is likely that corticosteroid therapy needs to be prescribed more than 5 days or until clinical improvement because of the risk of increased viral load that could lead to increased inflammation after the withdraw of corticosteroid therapy (14).

Several biological markers have been identified to modulate the course of COVID-19 (15). A recent meta-analysis showed that a decrease in lymphocytes count or lymphocyte to C-reactive protein ratio (LCR) discriminate severe cases of COVID-19 (16).

We designed a retrospective study to evaluate if early use of MP (7 days after COVID-19 symptoms first appeared) and for a longer duration is associated to a reduced risk of death or admission in intensive care unit (ICU), for patients with severe COVID-19 infection, hospitalized in Pulmonary or Cardiology Department of the Groupe Hospitalier de la Région de Mulhouse et Sud Alsace (GHRMSA), Mulhouse, France.

After collegial discussion between pneumologist and ICU staff and according to an internal protocol, 23 patients were treated by MP 1-2 mg/kg/day during 7 days followed by prednisolone at a gradually reduced dosage, for 4 to 6 weeks. The corticosteroid treatment was started one week after COVID-19 symptoms first appeared. All cases of COVID-19 patients that required oxygen were treated with pre-emptive intravenous cefotaxime (1g, 3 times per day for 7 days).

The criteria for ICU admission/intubation were worsening of respiratory failure despite maximal oxygen therapy, hemodynamic instability and neurological deterioration.

We also evaluated if the normalization of lymphocyte count between day 1 and day 7 could predict the prognosis.

## Methods

This observational study was conducted in the Pulmonary and Cardiology department of a 2409-bed regional hospital (Mulhouse-Sud Alsace Hospital, France). This study complied with the Declaration of Helsinki and was approved by the Institutional Review Board of the French Learned Society for Respiratory Medicine – “Société de Pneumologie de Langue Française” (CEPRO 2020-055).

### Participants

255 patients were admitted in the Pulmonary and Cardiology department between March 14^th^ to June 5^th^ 2020, with confirmed SARS-COV2 infection and/or acute respiratory distress (bilateral pulmonary infiltrates on chest x-ray on CT-scan and need for standard oxygen therapy between 1L/min and 15L/min).

The exclusion criteria were: (1) absence of laboratory confirmation of SARS-COV-2 infection and absence of typical COVID-19 CT scan profile, (2) death in the first 48 hours after admission, (3) transfer to ICU in the first 48 hours after hospitalization (these patients did not have the opportunity of corticosteroids treatment).

Finally, 181 patients were included and retrospectively assigned to intervention group if they were treated by corticosteroids during hospital stay, or to the control group, otherwise.

### Procedures

Clinical data, including morphological characteristics, comorbidities, vital signs and provided treatments was manually collected. Laboratory data at admission and discharge was automatically extracted from our health information system. Data management was performed in a reproductible way.

### Outcomes

The primary outcome was ICU admission, or death, during hospitalisation. A “do not resuscitate” (DNR) decision has sometimes been made, after collegial discussion between pulmonologists and ICU staff at admission, taking into account the patient’s opinion, age and comorbidities. Secondary outcomes related to corticosteroid therapy included 28-day all-cause mortality. The relationship between the outcome and the correction of lymphopenia or evolution of lymphocytes to C-reactive protein ratio (LCR), between days 1 to 7, was studied for exploration purpose.

### Statistical analysis

Descriptive analysis was expressed as numbers and proportions for categorical variables, and as means and standard deviations or median and quartiles for quantitative variables with nearly normal or non-normal distribution, respectively. No particular handling of missing data was performed, and analyses were restricted to complete cases.

Bivariate comparisons of covariates between treated (corticosteroid therapy) and control group on the one hand, and relation between each covariates and outcomes in the other hand, were performed trough Chi square test for categorical covariates, and parametric (t test) or non-parametric (Wilcoxon) tests for quantitative ones, depending on their distribution.

To restore comparability between the two groups, a 1:3 nearest neighbour propensity score matching was conducted, based on three covariates: age, hypertension or diabetes comorbidity, and maximal dose of oxygen therapy in first 48 hours.

We stratified the severity of respiratory failure in three categories: severe pneumonia defined patients requiring more than 10L/min of oxygen flow, moderate pneumonia defined patients requiring between 5 and 10 L/min of oxygen flow and mild pneumonia defined patients requiring less than 5L/min of oxygen flow.

The primary outcome was first analysed using a conditional logistic regression model applied on the matched groups. Several sensitivity analyses were carried out: (1) logistic regression performed on the whole population adjusted on the propensity score, (2) ‘naive’ logistic regression on the whole population adjusted on the same covariates than those used for matching. Same analyses were used for 28-day mortality.

The evolution of lymphocytes count and lymphocytes to CRP ratio, between days 1 to 7, was evaluated for description purpose.

Analyses were performed using R version 3.5.2 for Linux 64 bits. All statistic tests were two-sided and a significance level of 0.05 was considered. Odds ratio were reported with their 95% CI.

## Results

### Population characteristics

Out of 255 patients, hospitalized in our departments between March 14^th^ to June 5^th^ 2020, 181 met the inclusion criteria, out of which 23 (12.7%) of them received corticosteroids during hospitalization. Figure 1 shows the study profile and the reasons for exclusion.

**Figure 1.**
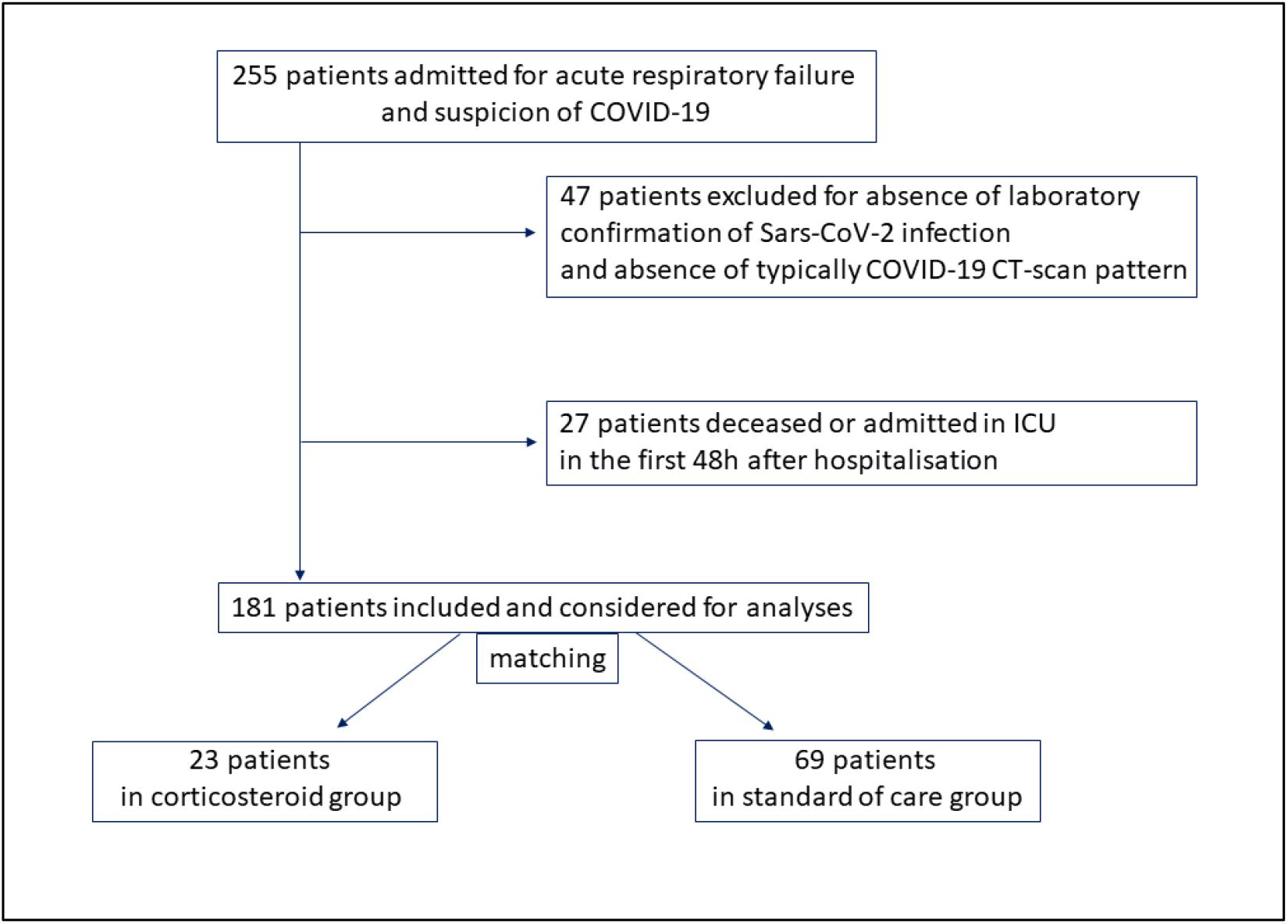
Study profile.

We found that the patients treated by corticosteroids had a more severe respiratory failure, they were more likely to have cardiovascular comorbidities and they received more likely a DNR decision. We did not find any differences in terms of age, sex, BMI, respiratory comorbidities, inflammatory status, lymphocyte count, Ddimers dosage, NT-proBNP or Troponin level or delay of COVID-19 symptoms (Table 1). The majority of patients (82.3%) had laboratory confirmation SARS-COV-2 infection. The remaining patients presented a typically COVID-19 CT scan profile.

**Table 1.**
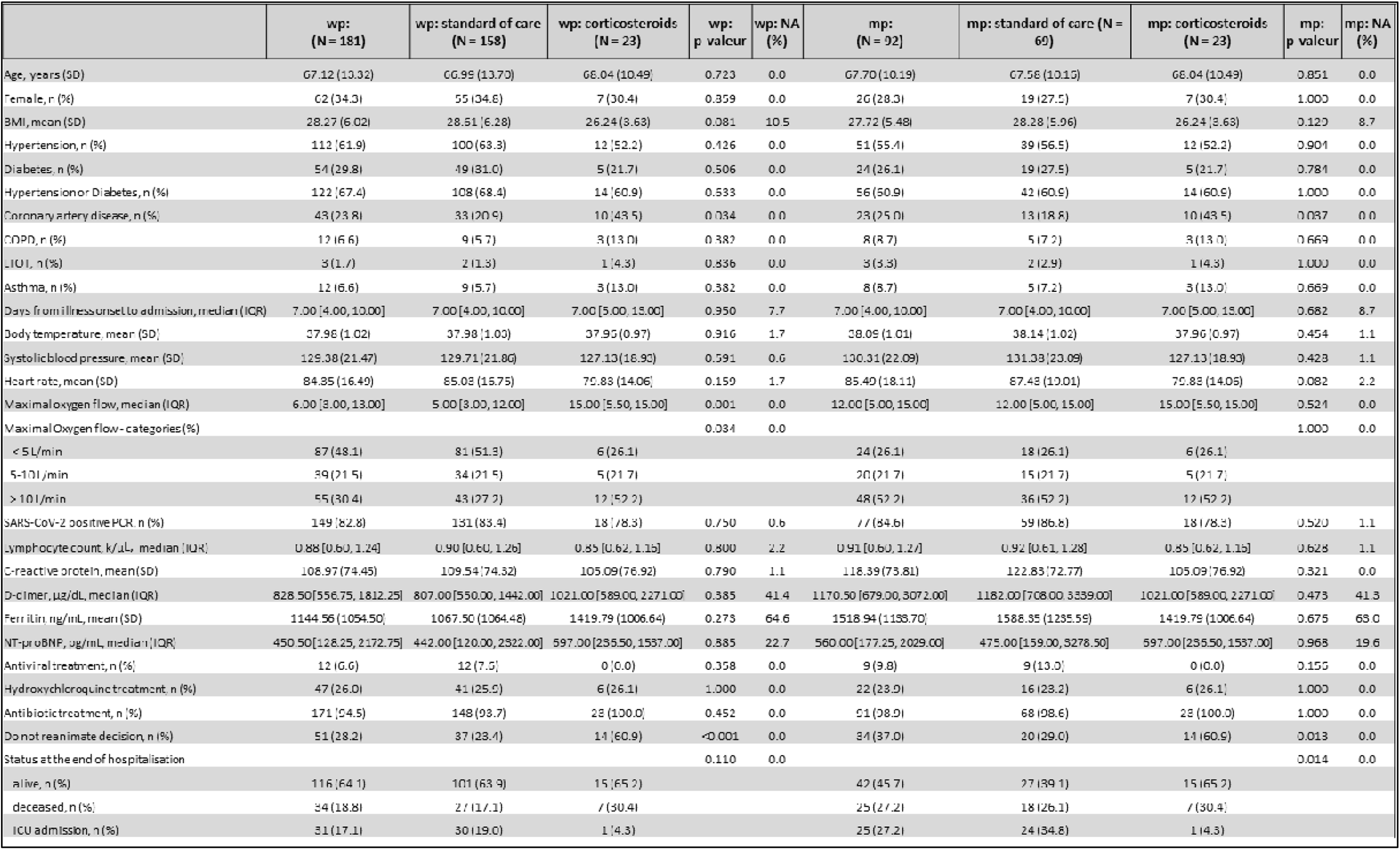
Patients characteristics Continuous, normally distributed variables, are presented as mean ± SD. Continuous, non-normally distributed variables are presented as median (interquartil e range). Categorial variables are presented as No. (%). Abbreviation s: wp, whole population; mp, matched population; BMI, body mass index; COPD, chronic obstructive pulmonary disease; LTOT, long-term oxygen therapy; PCR, polymerase chain reaction; ICU, intensive care unit; IQR, interquartile range; SD, standard deviation.

In order to evaluate the efficacy of corticosteroid therapy, we compared the intervention group (23 patients) to a control group (69 patients) adjusted in terms of age, degree of respiratory impairment and history of arterial hypertension and diabetes. The characteristics of the adjusted and non-adjusted study population are presented in Table 1. In the case of non-adjusted model (181 patients), no significant difference was seen between patients regarding the primary or secondary outcomes.

In the adjusted model (92 patients), 50 patients met the primary outcome of death or ICU admissions because of respiratory impairment (25 of whom died and 25 of whom were admitted in ICU). The overall composite outcome (death or ICU admission during hospitalisation) was 42/69 (60.9%) in the control group versus 8/23 (34.8%) in the group of patients treated by systemic corticosteroids (p= 0.018; odds ratio 0.24; 95% CI 0.07-0.79; with absolute risk of death or ICU admission reduced by 26% and relative risk of death or ICU admission reduced by 43%) (Figure 2). In a subgroup analysis, we found a trend to higher absolute benefit among patients with higher oxygen flow (reduction of composite outcome by 39% for patients requiring an oxygen flow higher than 10 L/min, by 13% for patients requiring an oxygen flow between 5 and 10L/min and 11% for patients requiring an oxygen flow less than 5L/min) and with higher inflammatory status (reduction of composite outcome by 42% if CRP level ≥ 100 mg/dL and by 7% if CRP level < 100mg/dL).

**Figure 2.**
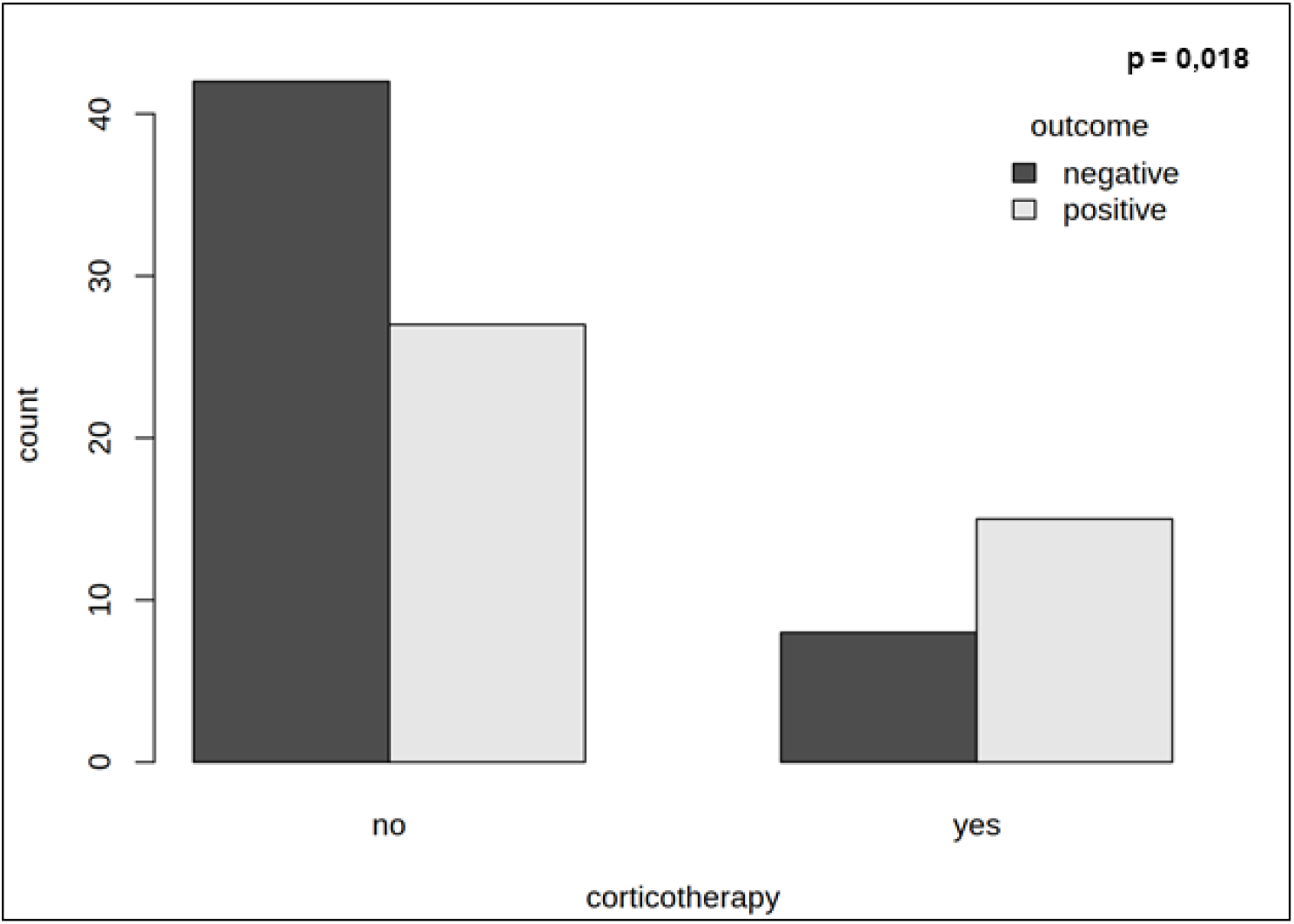
Comparison between groups in terms of primary composite outcome

Regarding secondary outcome, the 28-days mortality was not different between the two groups (31.9% in the control group vs. 30.4% in the interventional group, p=0.9). The correction of lymphopenia between day 1 to day 7 was associated to better composite outcome (p=0.006; odds ratio 0.17, 95% CI 0.05-0.61; with absolute risk reduction by 28% and relative risk reduction by 75%) (Table 2). Higher LCR at day 7 seemed to be associated with better outcome (Figure 3).

**Table 2.** Primary, secondary and exploratory outcomes

OR, odds ratio; CI, confidence interval
|  | Total<br>(N = 92) | standard of care (N = 69) | corticosteroids (N = 23) | OR (95% CI) | p-value |
| --- | --- | --- | --- | --- | --- |
| **Primary outcome** |  |  |  |  |  |
| Mortality and/or ICU transfer during hospitalisation, n/N | 50/92 | 42/69 | 8/23 | 0.24 (0.07 - 0.79) | 0.002 |
| **Secondary outcome** |  |  |  |  |  |
| 28-day mortality, n/N | 29/92 | 22/69 | 7/23 | 0.92 (0.3 - 2.84) | 0.87 |
|  | Total<br>(N = 95) | Normalization of lymphopenia<br>(N = 33) | Non-normalization of lymphopenia<br>(N = 62) | OR (95% CI) | p-value |
| **Exploratory outcome** |  |  |  |  |  |
| Mortality and/or ICU transfer during hospitalisation, n/N | 69/95 | 3/33 | 23/62 | 0.17 (0.05 - 0.61) | 0.004 |

**Figure 3.**
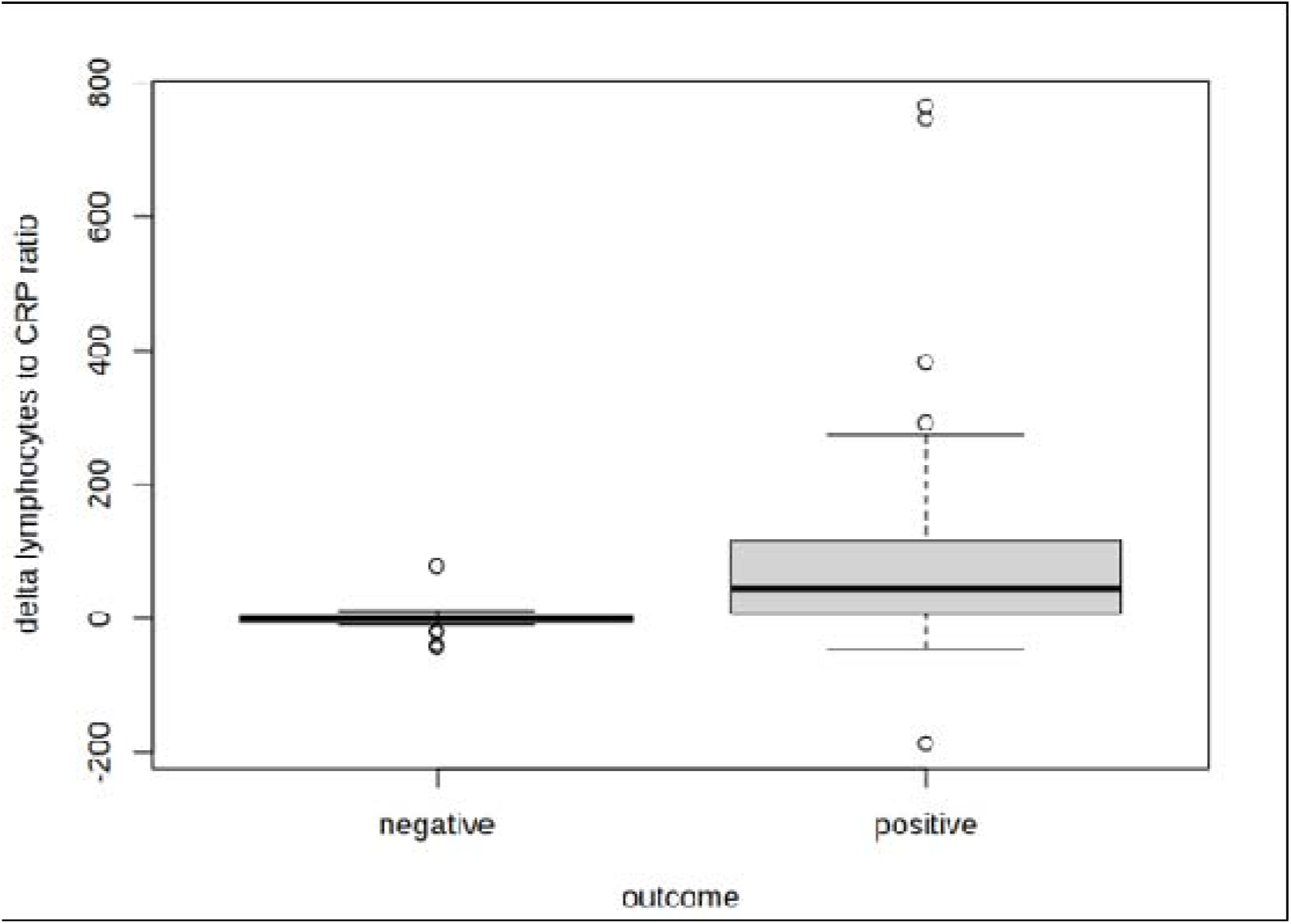
Exploratory outcome - evolution of lymphocytes to C-reactive protein ration, between day 1 to day 7. CRP; C-reactive protein.

## Discussion

The results of this study indicate that early treatment with MP, at 1-2 mg/kg/day during 7 days, followed by prednisolone at a gradually reduced dosage, for 4 to 6 weeks is associated with reduced mortality and fewer ICU admissions for worsening respiratory state. These findings are consistent for an adjusted population (by age, hypertension or diabetes comorbidity, and maximal dose of oxygen therapy during the first 48 hours). We used this corticosteroid therapy as a compensationate treatment for a majority of patients who received a DNR decision, based on patient’s wishes, age and comorbidities. Dexamethasone is now used commonly and considered as part of standard of care for oxygen requiring COVID-19 patients (9).

Less is known about the efficacy of other corticoid molecules in COVID-19 patients. The recently published METCOVID randomized trial showed that MP, at 1 mg/kg/day during 5 days, was not associated with lower rate of mortality in severe COVID-19 patients. One reason could be the shorter duration of corticoid treatment and a late start of MP therapy at an advanced stage of the disease. However, a subgroup analysis found a lower mortality rate for patients over 60 years old who received MP and these patients presented a higher level of inflammation status, with higher CRP levels (10).

In our study, we found a reduced rate of mortality and/or ICU admissions, with an absolute risk of composed outcome reduced by 26%. This result could be explained by the early start of corticosteroid therapy (during the second week of the disease) and the longer duration of treatment (4 to 6 weeks at a gradually reduced dose). A recent review of corticosteroid therapy studies showed a delayed viral clearance for SARS-CoV1 and MERS-CoV infections which can lead to more inflammation after the withdraw of corticosteroids (14).

The results from RECOVERY trial indicated a mortality benefit, mostly in severe cases of COVID-19 (9). Our study reaffirms these findings, with better results in case of patients receiving high oxygen flow and having a higher inflammatory status.

It is known that lymphopenia and low lymphocytes to C-reactive protein ratio at baseline could predict the severity of disease in patients with COVID-19 (16–18). We found that the correction of lymphopenia between day 1 to day 7 is associated to better prognosis and could be a predictive marker of better evolution of the disease. Secondly, higher lymphocytes to C-reactive protein ratio, between day 1 to day 7, seemed to be associated with better outcome.

Observational studies suggest a higher risk for secondary bacterial or fungal infections following the use of corticoids in viral infections (19,20). Our study was not designed to evaluate the risk of infection secondary to corticosteroid treatment. Although, all the patients included in our analysis were hospitalized, monitored and treated by ceftriaxone in association to macrolide. We did not noticed nay fungal or bacterial infection in our population of study.

This study had several limitations. Our study analyses retrospectively a small sample size and therefore it is subject to confounding and bias. Since this study was done in a single center, these results cannot be generalized. However, we tried to reduce the bias and confounders by restoring comparability between the two groups, by 1:3 nearest neighbour propensity score, based on three covariates: age, hypertension or diabetes comorbidity, and maximal dose of maximal oxygen therapy in first 48 hours. Moreover, the patients treated by MP were more likely to have cardiovascular comorbidities. During the first 2 months of pandemic in France, we used this corticosteroid regime as a compassionate use and therefore this analysis is not based on a randomized allocation of therapy. Therefore, these results need to be confirmed in a larger, prospective study.

Last, due to limited number of patients we could not assess the effect of corticosteroid treatment on the respiratory functional recovery.

In conclusion, the use of methylprednisolone, at 1-2 mg/kg/day during 7 days followed by prednisolone at a gradually reduced dosage for 4 to 6 weeks, seemed to improve the prognosis (the mortality and/or ICU transfer) in COVID-19 patients, hospitalized in a conventional medical ward. Our exploratory analysis showed that the correction of lymphopenia during the first week could be a prognostic marker of a better evolution of the disease.

## Data Availability

Data are available on request by contacting the corresponding author, George Calcaianu, at.

## Funding

This research received no specific grant from any funding agency in the public, commercial, or not-for-profit sectors.

## Conflict of interest

The author(s) declare that there is no conflict of interest.

## Acknowledgments

The authors thank to Clémence Perrin and Jérémie Henry for acquiring data.

Data are available on request by contacting the corresponding author, George Calcaianu, at.

## Notes

### Competing Interest Statement

The authors have declared no competing interest.

### Author Declarations

This study complied with the Declaration of Helsinki and was approved by the Institutional Review Board of the French Learned Society for Respiratory Medicine: Societe de Pneumologie de Langue Francaise (CEPRO 2020-055).

